# Cohort profile: Recruitment and retention in a prospective cohort of Canadian health care workers during the Covid-19 pandemic

**DOI:** 10.1101/2023.04.14.23288575

**Authors:** Nicola Cherry, Anil Adisesh, Igor Burstyn, Quentin Durand-Moreau, Jean-Michel Galarneau, France Labrèche, Shannon Ruzycki, Tanis Zadunayski

**Author notes:** **Corresponding author** Nicola Cherry, Division of Preventive Medicine, 5-22 University Terrace, 8303 112 St, Edmonton, Alberta, Canada T6G 2T4.

## Abstract

**Purpose:** Health care workers (HCWs) were recruited early in 2020 to chart effects on their health as the COVID-19 pandemic evolved. The aim was to identify modifiable workplace risk factors for infection and mental ill-health.

**Participants:** Participants were recruited from four Canadian provinces, physicians (MDs) in Alberta, British Columbia, Ontario and Quebec, registered nurses (RNs), licensed practical nurses (LPNs) and health care aides (HCAs) in Alberta and personal support workers (PSWs) in Ontario. Volunteers gave blood for serology testing before and after vaccination. Cases with COVID-19 were matched with up to 4 referents in a nested case-referent study.

**Findings to Date:** 4964/5130 (97%) of those recruited joined the longitudinal cohort: 1442 MDs, 3136 RNs, 71 LPNs, 235 PSWs, 80 HCAs. Overall, 3812 (77%) were from Alberta. Pre-pandemic risk factors for mental ill-health and respiratory illness differed markedly by occupation. Participants completed questionnaires at recruitment, fall 2020, spring 2021, and spring 2022. By the 4^th^ contact, 127 had retired, moved away or died, for a response rate of 89% (4299/4837). 4567/4864 (92%) received at least one vaccine shot: 2752/4567 (60%) gave post-vaccine blood samples. Ease of accessing blood collection sites was a strong determinant of participation. Among 533 cases and 1697 referents recruited to the nested case-referent study, risk of infection at work decreased with widespread vaccination.

**Future Plans:** Serology results (concentration of immunoglobulin G (IgG)) together with demographic data will be entered into the publicly accessible database compiled by the Canadian Immunology Task Force. Linkage with provincial administrative health databases will permit case validation, investigation of longer-term sequalae of infection and comparison with community controls. Analysis of the existing dataset will concentrate on effects on IgG of medical condition, medications and stage of pregnancy, and the role of occupational exposures and supports on mental health during the pandemic.

**Strengths and limitations:**

- Recruitment of a broad spectrum of health care workers close to the start of the COVID-19 pandemic through their professional organizations
- Consent to link to records held by public health departments allows for validation of self-reports of vaccinations and episodes of COVID-19 infection
- Repeated contacts permit charting the evolution of anxiety, depression and sources of stress through the course of the pandemic
- The inclusion of a nested case-referent study allows exposure reporting in near real time
- The absence of good denominator data limits the ability to examine recruitment bias

N=4216

## Introduction

This paper reports on the establishment of a longitudinal cohort of health care workers (HCWs) in different settings (hospitals, residential institutions, the community) and including workers with both specialist training in medicine and healthcare and those working at the front line of patient care but with training more tailored to immediate needs. The aim was to establish a prospective cohort to examine modifiable factors in the work place that might serve to mitigate risk of either infection or mental distress. Two additional study groups were later identified within the cohort, a serology sub-cohort to address immune response to vaccination and infection and a nested case-referent study to obtain near real time data on occupational exposures related to infection.

Longitudinal studies are prone to non-response bias if the attrition between recruitment and final follow up is related to the outcomes of interest. We aimed to keep attrition to a minimum and report here on factors systematically related to non-response.

### Cohort description

#### Recruitment procedures

Cohort recruitment began in Alberta in March 2020 and was extended to include HCWs in the four largest provinces of Canada (Alberta, British Columbia (BC), Ontario and Quebec). It comprised physicians (MDs), registered nurses and psychiatric nurses (RNs) licensed practical nurses (LPNs), personal support workers (PSW) and health care aides (HCAs). Potential participants were contacted, where possible, through their professional or workplace organization.

MDs in Alberta, BC and Ontario were approached through their provincial college, the licensing authority for physicians. In Alberta and BC, the colleges forwarded recruitment materials, explaining the aims of the study, to physicians who contacted the research team if they wished to take part. In Ontario the college provided contact emails to the study team to contact physicians directly but the college requested the emails were not used for further follow-up: physicians who had responded to the initial mailing providing contact details were retained. The Collège des Médecins du Québec agreed to provide links to the study on their website and requests were sent to the presidents of medical specialty federations to invite participation and posted on social media addressed to MDs. Only specialists in internal medicine, infectious disease, medical microbiology, respiratory medicine, critical care, anesthesiology, emergency medicine, geriatrics/care of the elderly, occupational medicine, public health and preventive medicine were approached together with physicians in family medicine and general practice. In BC and Ontario, to keep numbers manageable, community-based physicians were approached only from selected administrative areas (Hamilton, Peel. York and Ottawa in Ontario: Abbotsford, Burnaby, Chilliwack, Coquitlam, Delta, New Westminster, Port Coquitlam, Port Moody, Surrey and White Rock in BC).

RNs, LPNs and HCAs were recruited only in Alberta. RNs were recruited using e-mail addresses provided by the College and Association of Registered Nurses of Alberta (CARNA): the list of addresses only included those who had previously given approval to be approached about future (unspecified) research. Registered psychiatric nurses and LPNs were approached through their colleges, which sent out recruitment materials. The Alberta Register of Health Care Aides was held by the College of Licensed Practical Nurses of Alberta, which agreed to contact this group also. PSWs in Ontario were approached through the Ontario Personal Support Workers Association and through one of the larger groups providing home care outside the public system.

The recruitment information sent to potential participants included an introduction to the purpose of the study, together with links to a more detailed information sheet, to an online consent module and, if consent given, to the Phase 1 questionnaire (Supplementary Materials 1). The consent module asked also for consent to contact them again, together with space to give contact details (email and phone number). In Alberta, physicians and nurses who completed the consent but not the full questionnaire were invited to complete a short questionnaire to establish their demographic details. Participants in Alberta were also asked for consent to link their identity to the provincial administrative health database.

#### Structure and timelines

The full cohort was contacted to complete questionnaires at each of the four phases of the study. Phase 1 (baseline) from April 2020, Phase 2 in October 2020, Phase 3 in May 2021 and Phase 4 in May 2022. Cohort members were asked to give a pre-vaccine blood sample for serology testing in October 2020. Those who were willing and able to give serial blood samples post-vaccine gave such samples from March 2021-July 2022. A nested case-referent study recruited cases and matched referents from October 2020 to March 2022.

Newsletters were sent to update cohort members on study progress and preliminary findings in March 2021, November 2021 and March 2022.

#### Collection of information by questionnaire

Information was collected online using the Qualtrics survey platform, offered in both English and French. Reminders to complete were sent by email, with the alternative of a telephone interview offered to those not initially responding online. Where the participant had provided a phone number, this was used, where appropriate, to obtain information or as a reminder that a questionnaire had not yet been completed. At each phase subsequent to the baseline contact, a short questionnaire was offered in the weeks immediately prior to closing down on-line access to the questionnaire allowing those busy or reluctant to contribute core information. A payment of $50 (Canadian) was offered to those initially unwilling to complete the Phase 4, questionnaire. No other financial incentive was offered.

The questionnaires at the four phases had much information in common, including questions about work with COVID-19 patients, availability of personal protective equipment (PPE), infection with the COVID-19 virus and (after December 2020) vaccination. The Hospital Anxiety and Depression scale (HADS) [1] was included at each phase, together with a community acquired pneumonia questionnaire [2] and substance use (alcohol, tobacco, cannabis and medications for anxiety and sleep). All four questionnaires asked participants to rate statements about changes at work, confidence in working with infected patients, sources of worries and support and also asked an open-ended question on stressful events.The baseline (Phase 1) questionnaire included also questions on heath in the 12 months before the start of the pandemic (treatment for anxiety or depression, and use of medication for asthma), histories of tobacco use and chronic lung disease (bronchitis, emphysema or chronic obstructive lung disease). These were added to the questionnaire after the early recruitment of MDs who completed these questions at Phase 2. Age, gender, marital status and children under 18 years in the home were asked only at baseline: ethnicity was collected at Phase 2. The contact at Phases 2-4 contained questions on institutional supports for mental health and the phase 4, contact about lingering sequalae of a participant’s COVID-19 infection.

The questionnaires were adapted slightly to address each occupational group but were essentially the same for all. Phase 2,3 and 4 questionnaires were sent to everyone who had consented to follow-up unless they indicated they were unwilling to take part further (refused) had moved away from the participating provinces with no intent to return (moved away), had left healthcare work (either from choice or retirement), or were lost to trace (that is, neither email nor phone number were in service). Where a respondent indicated that they had some personal difficulty (such as illness or marital breakdown) that might not exclude them permanently, one or more follow-up contacts were omitted.

#### Collection of pre-vaccine serology samples

At the Phase 2 questionnaire (October 2020), participants were asked for permission to approach them to give a blood sample to allow analysis for serological markers of infection and immune response to vaccination and infection. It was not possible for the team to collect blood samples ‘in house’ but arrangements were made in each province for one or more clinical laboratories to take the sample, when approached by the participant with a study requisition. Although in Alberta the clinical laboratories covered the whole of the province, this was not the case elsewhere. In BC the provincial health care laboratories provided back-up in areas not covered by the commissioned commercial laboratory. In Ontario and Quebec, only commercial laboratories with limited coverage were available to collect samples: in those provinces participation in the serology study was effectively impossible for those living at a great distance from a commercial collection centre. Attempts to collect samples more informally and to transport them by courier to the commercial laboratory were not always successful, but a small number of participants in Ontario and Quebec were able to use this route.

#### Collection of post-vaccine serology samples

Those who had agreed to give a pre-vaccine serology sample were approached again in early 2021 to ask if they would be willing to take part in a serology sub-study giving samples at 4, 7 and 13 months after their first dose of vaccine: a 10-month sample was added subsequently, following the early observation of a rapid decline in antibody concentration between 4 and 7 months. Participants were asked to consider whether such a regimen of repeated samples would be feasible for them, in light of their distance from a collection centre and their personal commitments. Those who agreed were sent a requisition for each sample as it became due and asked to arrange for sample collection locally.

#### Nested case-referent study

A nested case-referent study was established within the cohort, following completion of the Phase 2 questionnaire, starting in October 2020 and continuing to 31^st^ March 2022. It considered exposures in the 21 days prior to a positive polymerase chain reaction (PCR) test. Those with a positive PCR test were then matched, as referents, with up to four people in the same province, on job classification, self-reported gender, and first vaccination date. Both cases and referents were asked, in the days immediately following case infection, to complete a detailed exposure questionnaire about experiences and exposures in the days leading up to the case’s positive test. More details are given in Cherry et al [3].

#### Updating of data on infection and vaccination

Each of the four questionnaires sent to the full cohort asked about positive tests for the SARS-CoV-2 virus and questionnaires 2-4 about vaccination shots received, once this became available to HCWs in mid-December 2020. In addition, participants were regularly reminded to contact the research team if they had a positive COVID-19 test or received a vaccine. Those in the post-vaccine sub-cohort were asked to complete a brief questionnaire at the time of each sample updating infection and vaccination status at the time of giving the sample. Participants were also asked for consent to approach the public health authorities in their province to obtain records of positive PCR tests and vaccination.

#### Protocol for reminders

At recruitment potential participants who were approached through their professional associations in Alberta and BC received two email reminders following the initial invitation. At each of the full cohort questionnaires post-recruitment non-responders received two email reminders (three if there was no telephone contact) and, if they had volunteered a telephone contact, up to two phone reminders. The same protocol was used for the case-referent study. Contact with the serology cohort was two-way with participants requesting help in arranging blood sampling and providing information on when and where samples had been collected. Those in this sub-cohort received reminders to book an appointment before a sample was due, the requisition before the due date and reminders to give a sample once the due date had passed. Because of the high engagement of participants, the overlapping of sub-cohorts and a policy of rapid personalized response to email or telephone queries, interaction between survey staff and cohort members became more informal and frequent than is implied by the reminder protocol outlined.

#### Numbers recruited

5130 HCWs gave consent for completion of the baseline questionnaire. Of these 4964 (96.8%) agreed to follow-up. The follow-up cohort comprised, by order of recruitment, 1442/1490 (96.8%) MDs, 3136/3227 (97.2%) RNs, 71/74 (95.9%) LPNs, 235/257 (91.4%) PSWs and 80/82 (97.6%) HCAs. Those who declined follow-up were more likely to be younger and male.

The estimated proportion accepting the invitation to join the study ranged from 18% of Alberta nurses to less than 4% of Ontario physicians, where recruitment could not continue beyond an initial email. In Alberta and BC about 7% of physicians approached joined the study, based on the number of emails sent. Estimates of the proportion participating were unsatisfactory for LPNs, HCAs and PSWs: where the numbers approached by email could not be ascertained and where, for HCAs, at least, emails were not actively maintained. In Quebec, recruitment of physicians depended on their response to a link on the College of Physician’s website. It seems likely that response to individual emails (had this been possible) would have been higher. We know nothing, other than professional role and province, about any of those not accepting the invitation to join and cannot investigate response bias from low response at recruitment.

#### Retention in the main cohort

By the time of the phase 4 questionnaire in spring 2022, 127 had become ineligible (2 were known to have died, 36 had moved from the study provinces and 89 had retired/left health care work), leaving a denominator of 4837. Among the 538 who remained in the cohort but who did not respond, 100 refused and contact details failed for 18. The research team made some contact with 61 of 420 remaining participants but the reason for eventual non-response was not known. The response rate at phase 4 was thus 88.9% (4299/4837) with 4731 completing a questionnaire at Phase 2 and 4519 at Phase 3.

Figure 1 gives the flow chart for recruitment and retention for the 4 phases of the main cohort.

**Figure 1:**
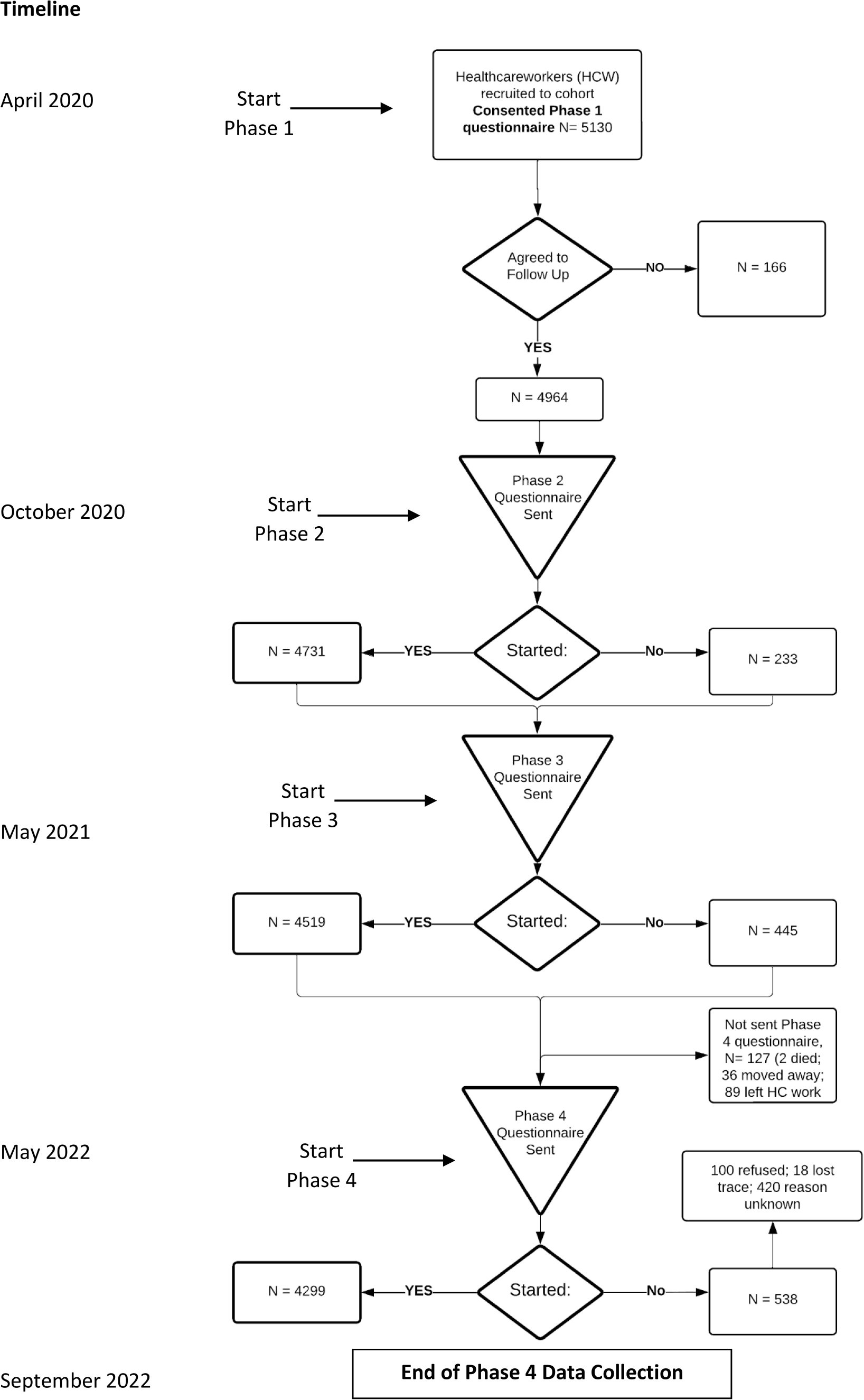
Flow chart of recruitment and retention for all phases of the main cohort

#### Characteristics of participants

At baseline, the majority of participants were working in Alberta (77%) and were aged 40 years or older (62%): PSWs (76%) and HCAs (73%) had a greater proportion over 40 yrs. Most were married or living as married (75%): fewer than half (43%) had a child aged <18 years living in their home. The majority of the participants were female (4111) with 848 male and five who reported their gender as non-binary. Overall, 83% reported their ethnicity as white, with the highest proportion in RNs (87%). Only 2% reported themselves as Indigenous and 1% as Black. (Table 1).

**Table 1:** Demographics of participants by work role*

|  | MD |  | RN |  | LPN |  | PSW |  | HCA |  | All |  |
| --- | --- | --- | --- | --- | --- | --- | --- | --- | --- | --- | --- | --- |
| Province | N | % | N | % | N | % | N | % | N | % | N | % |
| Alberta | 525 | 36.4 | 3136 | 100.0 | 71 | 100.0 | ? | ? | 80 | 100.0 | 3812 | 76.8 |
| British Columbia | 263 | 18.2 | ? | ? | ? | ? | ? | ? | ? | ? | 263 | 5.3 |
| Ontario | 513 | 35.6 | ? | ? | ? | ? | 235 | 100.0 | ? | ? | 748 | 15.1 |
| Quebec | 141 | 9.8 | ? | ? | ? | ? | ? | ? | ? | ? | 141 | 2.8 |
| Self-reported gender |  |  |  |  |  |  |  |  |  |  |  |  |
| Female | 805 | 55.8 | 2949 | 94.0 | 67 | 94.4 | 215 | 91.5 | 75 | 93.8 | 4111 | 82.8 |
| Male | 637 | 44.2 | 182 | 5.8 | 4 | 5.6 | 20 | 8.5 | 5 | 6.3 | 848 | 17.1 |
| Non-binary | 0 | 0.0 | 5 | 0.2 | 0 | 0.0 | 0 | 0.0 | 0 | 0.0 | 5 | 0.1 |
| Age at March 2020 |  |  |  |  |  |  |  |  |  |  |  |  |
| < 40 y | 464 | 32.2 | 1288 | 41.1 | 33 | 46.5 | 56 | 23.8 | 22 | 27.5 | 1863 | 37.5 |
| 40 y < 55 y | 543 | 37.7 | 1125 | 35.9 | 27 | 38.0 | 104 | 44.3 | 38 | 47.5 | 1837 | 37.0 |
| 55 y or older | 434 | 30.1 | 723 | 23.1 | 11 | 15.5 | 74 | 31.5 | 20 | 25.0 | 1262 | 25.4 |
| Age unknown | 1 | 0.1 | 0 | 0.0 | 0 | 0.0 | 1 | 0.4 | 0 | 0.0 | 2 | 0.0 |
| Married or living as married | 1158 | 81.1 | 2238 | 74.2 | 46 | 64.8 | 141 | 61.3 | 40 | 50.0 | 3623 | 75.1 |
| Children <18 y at home | 689 | 48.0 | 1292 | 41.3 | 23 | 32.4 | 80 | 34.5 | 23 | 28.7 | 2107 | 42.6 |
| Ethnicity |  |  |  |  |  |  |  |  |  |  |  |  |
| Indigenous | 5 | 0.4 | 69 | 2.4 | 5 | 8.3 | 2 | 1.1 | 2 | 2.8 | 83 | 1.8 |
| Asian | 184 | 13.6 | 191 | 6.6 | 4 | 6.7 | 12 | 6.7 | 9 | 12.7 | 400 | 8.8 |
| Black | 15 | 1.1 | 28 | 1.0 | 0 | 0.0 | 9 | 5.1 | 4 | 5.6 | 56 | 1.2 |
| White | 1052 | 78.0 | 2488 | 86.5 | 49 | 81.7 | 140 | 78.7 | 54 | 76.1 | 3783 | 83.4 |
| Other | 93 | 6.9 | 101 | 3.5 | 2 | 3.3 | 15 | 8.4 | 2 | 2.8 | 213 | 4.7 |
| N | 1442 |  | 3136 |  | 71 |  | 235 |  | 80 |  | 4964 |  |

\* MD, medical doctor; RN registered nurse or registered psychiatric nurse; LPN, licensed practical nurse; PSW, personal support worker; HCA, health care aide.

At Phase 1 (baseline) 88% (4184/4760) of those known to be working were in direct contact with patients (either ‘hands on’ or virtually), and of these 58% had all or part of their work in a hospital setting, 35% in the community and 12% in a residential setting. A small proportion (7%) worked only in another setting, including roles related to COVID-19 (screening, contact tracing). Physicians (65%) and nurses (61%) were most likely to be working in a hospital setting, whereas PSWs and HCAs were more likely to be working in the community (including the client’s home) or in a residential community such as a care-home (Table 2).

**Table 2:** Workplace setting* at first contact in 2020 by work role**

|  | MD |  | RN |  | LPN |  | PSW |  | HCA |  | All |  |
| --- | --- | --- | --- | --- | --- | --- | --- | --- | --- | --- | --- | --- |
|  | N | % | N | % | N | % | N | % | N | % | N | % |
| Participant has one-on-one contact with patients |  |  |  |  |  |  |  |  |  |  |  |  |
| Hospital setting | 834 | 65.0 | 1555 | 60.6 | 35 | 54.7 | 5 | 2.5 | 13 | 18.3 | 2442 | 58.4 |
| Community setting | 585 | 45.6 | 690 | 26.9 | 18 | 28.1 | 139 | 68.8 | 30 | 42.3 | 1462 | 34.9 |
| Residential setting (e.g. care home) | 128 | 10.0 | 238 | 9.3 | 13 | 20.3 | 73 | 36.1 | 35 | 49.3 | 487 | 11.6 |
| Other setting with patient contact only | 61 | 4.8 | 212 | 8.3 | 3 | 4.7 | 10 | 5.0 | 4 | 5.6 | 290 | 6.9 |
| N | 1283 |  | 2564 |  | 64 |  | 202 |  | 71 |  | 4184 |  |
| No one-on-one contact with patients |  |  |  |  |  |  |  |  |  |  |  |  |
| Public health | 17 | 15.7 | 57 | 12.7 | 1 | 33.3 | 4 | 40.0 | 3 | 50.0 | 82 | 14.2 |
| Administration | 18 | 16.7 | 113 | 25.2 | 1 | 33.3 | 1 | 10.0 | 0 | 0.0 | 133 | 23.1 |
| Teaching/research | 15 | 13.9 | 86 | 19.2 | 1 | 33.3 | 0 | 0.0 | 0 | 0.0 | 102 | 17.7 |
| Other setting with no patient contact only | 67 | 62.0 | 209 | 46.5 | 1 | 33.3 | 5 | 50.0 | 3 | 50.0 | 285 | 49.5 |
| N | 108 |  | 449 |  | 3 |  | 10 |  | 6 |  | 576 |  |
| No information on work settings |  |  |  |  |  |  |  |  |  |  |  |  |
| Not worked: all reasons | 23 |  | 43 |  | 1 |  | 3 |  | 3 |  | 73 |  |
| Working: details unknown | 20 |  | 66 |  | 3 |  | 15 |  | 0 |  | 104 |  |
| Unknown if working | 8 |  | 14 |  | 0 |  | 5 |  | 0 |  | 27 |  |
| N | 51 |  | 123 |  | 4 |  | 23 |  | 3 |  | 204 |  |
| Total N | 1442 |  | 3136 |  | 71 |  | 235 |  | 80 |  | 4964 |  |

\*Respondent can report working in multiple setting.
\*\*MD, medical doctor; RN registered nurse or registered psychiatric nurse; LPN, licensed practical nurse; PSW, personal support worker; HCA, health care aide.

Smoking habit and health in the 12 months before the start of the pandemic differed markedly with work role. Physicians (1%), RNs (6%) and LPNs (13%) reported less often than PSWs (32%) or HCAs (25%) that they had smoked tobacco in the 12 months before March 2020 and were less likely to report chronic lung disease other than asthma. Overall, 24% reported having been treated for anxiety or depression in the 12 months before the pandemic. MDs were least (17%) and LPNs most (48%) likely to report treatment for these conditions (Table 3).

**Table 3:**
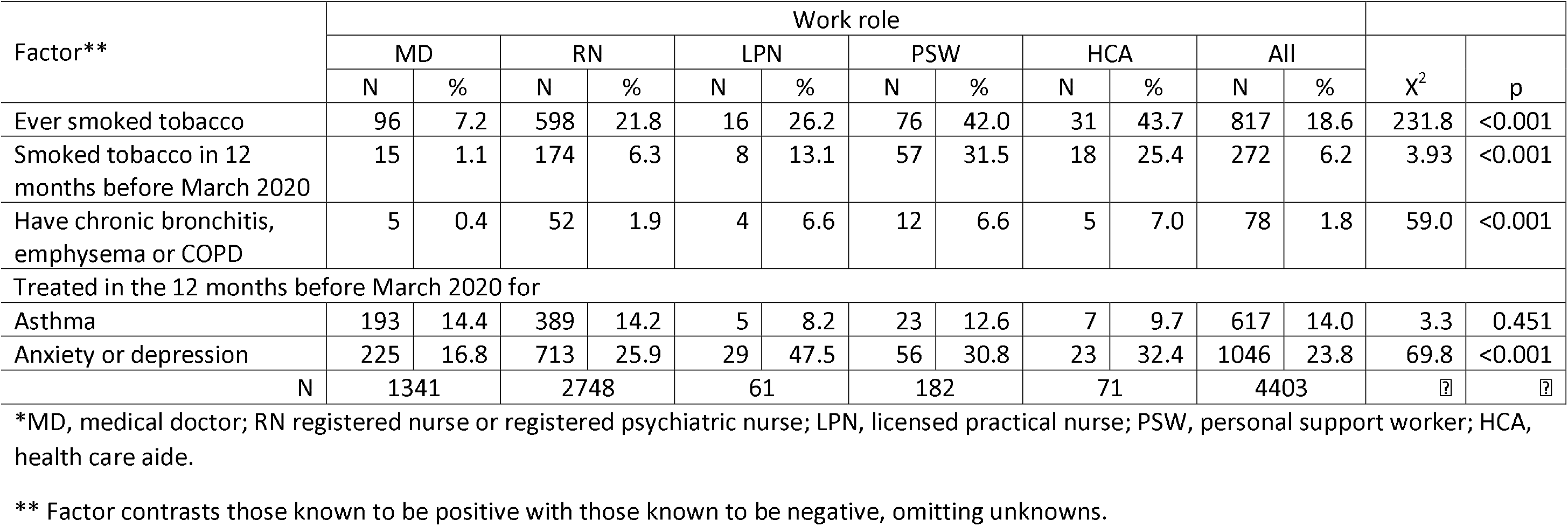
Smoking and health in the 12 months before the start of the pandemic by work role*

Factors associated with attrition between baseline and Phase 4 are shown in Table 4, with completion tabulated by data collected at baseline. Partial completion of the baseline questionnaire was a strong predictor of attrition, but the Phase 4 completion rate was 79% even in this group. As seen in the bivariate logistic regression in Table 4, completion was higher with greater age and in those married or living as married and lower in those smoking in the year before the start of the pandemic. The missing data on smoking, lung disease, use of asthma medication, anxiety and depression seen in the bivariate analysis arose from a failure to complete the full baseline questionnaire and was strongly related to attrition at Phase 4. The multivariable analysis presented in Table 4 is limited to those fully completing the baseline questionnaire. It supported the bivariate results for all factors other than smoking, where the effect was less marked. Treatment for anxiety or depression in the 12 months before the start of pandemic showed only a slight association with attrition in the bivariate analysis: this was reduced in the multivariable model.

**Table 4:**
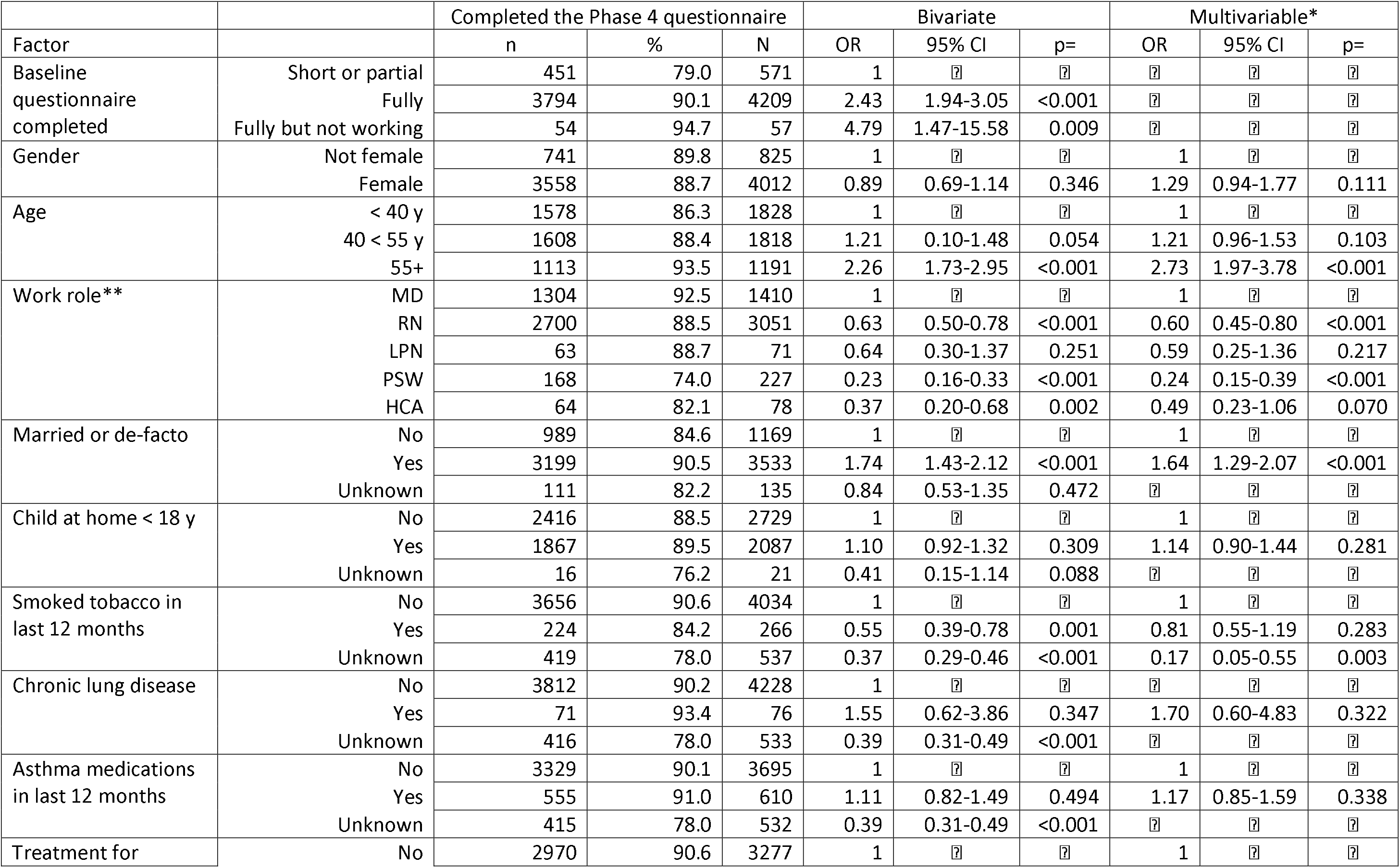
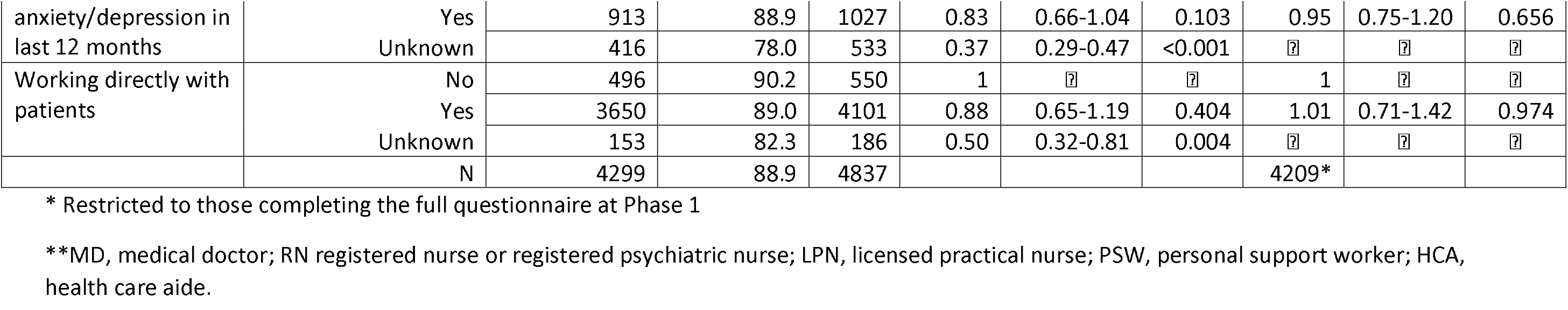
Completion of the Phase 4 questionnaire in Spring/Summer 2022. Participants eligible to receive a Phase 4 questionnaire only.

#### Participation in the serology study

The numbers giving serology samples pre- and post-vaccine are given in Table 5: 59% (2940/4964) of the full cohort gave a blood sample for serology before vaccination and 60% (2752/4567) at least one sample post-vaccine (excluding the 397, 8.0%, not known to have been vaccinated). Participation was much lower in PSWs than in other HCWs with only 16% giving a pre-vaccine sample and 12% post-vaccine. Factors associated with giving pre and post-vaccine serology samples are shown in Supplementary Materials 2. Women, older HCWs, nurses, those who were married and those working directly with COVID-19 patients were more likely to give a sample. Smokers and those having a child in the home were less likely to give a pre-vaccine sample (Table ST1). MDs and RNs were most likely to join the post-vaccine serology sub-cohort, as were those who were older and married (Table ST2). Having a child at home and smoking were again associated with a lower participation.

**Table 5:** Blood samples for serology pre and post vaccine by work role* *MD, medical doctor; RN registered nurse or registered psychiatric nurse; LPN, licensed practical nurse; PSW, personal support worker; HCA, health care aide. ** among those reporting at least one vaccine dose.

|  | MD |  | RN |  | LPN |  | PSW |  | HCA |  | All |  | X<sup>2</sup> | p= |
| --- | --- | --- | --- | --- | --- | --- | --- | --- | --- | --- | --- | --- | --- | --- |
|  | N | % | N | % | N | % | N | % | N | % | N | % |  |  |
| Pre-vaccine serology sample |  |  |  |  |  |  |  |  |  |  |  |  |  |  |
| No | 606 | 42.0 | 1143 | 36.4 | 33 | 46.5 | 197 | 83.8 | 45 | 56.3 | 2024 | 40.8 | 219.6 | <0.001 |
| Yes | 836 | 58.0 | 1993 | 63.6 | 38 | 53.5 | 38 | 16.2 | 35 | 43.8 | 2940 | 59.2 |  |  |
| Total | 1442 | 100.0 | 3136 | 100.0 | 71 | 100.0 | 235 | 100.0 | 80 | 100.0 | 4964 | 100.0 |  |  |
| At least one post-vaccine serology samples** |  |  |  |  |  |  |  |  |  |  |  |  |  |  |
| No | 535 | 38.9 | 1059 | 36.7 | 33 | 49.3 | 153 | 88.4 | 35 | 54.7 | 1815 | 39.7 | 199.0 | <0.001 |
| Yes | 839 | 61.1 | 1830 | 63.3 | 34 | 50.7 | 20 | 11.6 | 29 | 45.3 | 2752 | 60.3 |  |  |
| Total | 1374 | 100.0 | 2889 | 100.0 | 67 | 100.0 | 235 | 100.0 | 80 | 100.0 | 4567 | 100.0 |  |  |

#### Participation in the case-referent study

Of 542 cases eligible for the case-referent study,534 (99%) completed the case-referent questionnaire. Among 1815 selected referents 1697 (93.5%) completed [3].

#### Findings to date

As reported here, occupational groups differed markedly on pre-pandemic health indicators including smoking, chronic lung disease and treatment for anxiety and depression. Retention in the main cohort and recruitment to the serology sub-cohort was higher in older and married HCWs. In the case-referent study it was evident that unvaccinated HCW, in the early phase of the pandemic were at greater risk of infection if they worked hands-on with patients with Covid-19, on a ward designated for care of infected patients, or handled objects used by infected patients. Later in the pandemic, with almost universal HCW vaccination, risk from working with infected patients was much reduced [3].

## Discussion

The strengths of the cohort include baseline data collected in the first weeks of the pandemic from a spectrum of HCW roles and job settings. The great majority of those recruited have been retained, completing questionnaires throughout the pandemic. Participation in the serology cohort by 60% of the cohort was high, given the time and organization needed for up to four post-vaccine samples. It is a strength to be able to document factors associated with participation in the serology sub-cohort. The case-referent study, nested within the cohort, allowed near ‘real-time’ reporting of exposures. The study depended on self-reporting of infection and vaccination and the ability to check these reports against public health records in at least two provinces (Alberta and BC) gives strength, particularly to the analysis of determinants of antibody response. This established cohort, with repeated measures over the course of the pandemic, provides sound data for analysis of workplace and personal factors related to vaccination, infection with the SARS-CoV-2 virus and adverse health outcomes of COVID-19 illness, and of the evolution of mental ill-health during the pandemic.

The low response rate to the invitation to join the cohort was a weakness. Those recruited were potentially unrepresentative and this limits the generalizability of the results. Challenges in setting up the cohort included finding organizations with nominal lists and contact details who were willing and able, at the start of the pandemic, to supply those details or to act on behalf of the study to contact potential participants. Although considerable efforts were made to recruit and retain HCW other than physicians and nurses, workers in other roles were under-represented. A further weakness is that episodes of COVID-19 may have been under-reported and this cannot be checked in all provinces. Asymptomatic and unrecognized cases can be charted only for those giving serology samples (where nucleocapsid antibodies would result from infection rather than vaccination). Arrangements for collection of serology samples were not uniform across provinces. A high proportion of participants were from Alberta, resulting from the decision to limit recruitment of nurses to those working in that province, which reduces the capacity to generalize findings to nurses working across Canada: only MDs contributed data from all four participating provinces.

A major challenge for longitudinal studies is attrition between contacts [4]. This is particularly of concern if the reason for non-response is closely associated with the study outcome and risk factors of interest [5]. The attrition rate (11%) in the study reported here is lower than other population-based studies of HCWs during the pandemic, ranging from 45% [6] and 57% [7] to 68-69% [8] [9] over shorter periods of follow-up. Low attrition rates over multiple contacts are rare in community-based studies, although de Graaf et al [10] achieved a 20% attrition rate in the first follow-up in a prospective psychiatric epidemiological study. Nguyen et al [11] report an attrition rate of 15% in a community-based cohort of COVID-19 with follow-up of 16 months. Both De Graaf and Nguyen attributed their successful retention to the large time-investment to encourage respondents, as was done in the present study. Analysis of those not retained suggested that attrition was not random. As in other studies age, marriage and higher socioeconomic status (here represented by MDs and RNs) were associated with higher compliance and smoking with lower [12] [13] [14]. Importantly, given that a main focus of the study was mental health, the lack of an association between attrition and treatment for anxiety or depression in the 12 months before the pandemic was encouraging. Previous studies have differed on the importance of baseline mental health as a predictor of attrition, with some [10] [15] [16] reporting (as here) no relationship while others [6] [14] [17] [18] found those with mental ill-health were less likely to respond at follow-up. Despite high participation in the cohort during the pandemic, it will be important to remain vigilant of the impact attrition may have on causal inference.

Recruitment to the serology sub-sample was not random, not least because of the less satisfactory arrangements for sample collection in Ontario and Quebec. The rate in PSWs in Ontario (16%) for giving a pre-vaccine sample was less than half that for HCAs in Alberta (44%) although these groups were similar in other ways. Inspection of pre-vaccine samples within physicians showed samples from only 40% of MDs in Ontario and Quebec but 78% for Alberta (where collection sites covered the whole province) and 62% for BC where black holes for collection were fewer than in Ontario and Quebec. Similar, though more marked, differences were evident for post-vaccine samples, where commitment was asked for repeated blood samples over nine months: here rates for PSWs were only 12% compared with 45% for HCAs, and Ontario MDs with the lowest rate of participation among MDs at 44%. Bias in participation may be of limited importance for a serology study but biased estimates of IgG concentration in HCWs could result if rurality or remoteness was differentially associated both with infection or vaccination and difficulty in giving blood samples

### Future plans

Results of the serology analysis (the concentration of IgG against the receptor binding site of the spike protein of the SARS-CoV-2 virus) will be entered, together with demographic data, into the publicly accessible database being compiled by the Canadian Immunology Task Force [19]. Further, we will take advantage of the data available through Canada’s public health care system. Using these administrative data, we will investigate longer term sequalae of COVID-19 infection, and the influence of pre-existing health conditions by linking those in the Alberta cohort to administrative health records both before and since the start of the pandemic. We have in place the necessary consents and approvals to do this. We will validate self-reports on infection and vaccination as these health reports become available from cooperating provinces. In Alberta we have in place an agreement to link administrative health records of each HCW to the health records of 5 community referents, matched on age, sex and geographic region to allow comparison of infection rates in the cohort. As further collaborations we are exploring comparing data from this cohort with a parallel cohort of paramedics recruited from 5 Canadian provinces including Alberta, British Columbia and Ontario [20].

Within data currently available on the present cohort, serology analysis concentrates on the extent to which medical conditions, medications and stage of pregnancy add to vaccination, infection, and age in determining IgG. Mental health changes during the pandemic are documented through scores on the Hospital Anxiety and Depression Scale (HADS) completed at each of 4 contacts, with changes during the pandemic examined by work demands, availability of PPE, vaccination and infection, within risk groups defined by pre-pandemic health indicators. An analysis of stressful events reported by participants will describe the evolution of sources of stress since the start of the pandemic. The extent to which supports offered through work modify anxiety and depression scores is being assessed. The severity of response to infection is considered in linked analyses of symptom reporting, time off work, and post-COVID-19 symptoms.

### Patient and Public Involvement

The first phase of the study was set up very rapidly after Covid-19 was reported in Canada. Discussions between the study team and leadership of health care worker organizations in the four provinces were arranged to ensure the approach taken and study questions were acceptable to participants, and to inform recruitment strategies. We solicited feedback about the research topics throughout Phase 1, and Phase 2 was designed taking account of feedback on the initial questionnaire and the perceived need by the leaders of participant healthcare organizations for additional information on mental health supports. Ongoing communication towards participants was individual and frequent. Participants received three newsletters with updates on preliminary results and further explanatory background information, with options to comment on the study and to submit questions. Participants in the serology sub-study received their individual post-vaccine antibody results.

## Data Availability

The data on which this manuscript is based will be made available on reasonable request to the corresponding author. The data are subject to a data sharing agreement with the major funder, the Canadian Immunology Task Force. Through the databank currently being built individual anonymized data will be made available to researchers internationally on request to the CITF.

## Author Contributions

NC, AA, IB, QD-M, J-MG, FL and SR contributed to the initial design of the study. J-MG established the dataset. TZ coordinated the ongoing acquisition of data. NC analyzed the data in discussion with IB and J-MG. NC drafted the manuscript. All authors contributed to the interpretation of the data and critically revised the draft. All read and approved the final manuscript

## Statement of Ethics Approval

Approval for each element of the study was given by the University of Alberta Heath Ethics Board (Pr000099700). The study was also reviewed and approved by Unity Health Toronto Research Ethics Board (REB# 20-298) for those elements coordinated locally for the Ontario participants. The research was carried out in accordance with the Declaration of Helsinki. All participants gave online written informed consent

