## Supplementary Materials for "Cohort profile: Recruitment and retention in a prospective cohort of Canadian health care workers during the Covid-19 pandemic"

Supplementary materials 1

Baseline (Phase 1) questionnaire.

Example given is for health care aides in Alberta.

Health Care Aides from Alberta - May 2020

Start of Block: Introduction

Intro **The impact of Covid-19 on the health of Alberta Health Care Aides
 Online consent and survey** Thank you for considering joining this study. Before completing the questionnaire you need to read the [information sheet](https://sites.ualberta.ca/~covidhca/info.html) and sign the consents. If you have any questions you can phone us on 1–866–492–6093 or contact us by email at. We will get back to you very rapidly.

 It is entirely your decision whether or not you take part in the study. If you do decide to take part, please be assured we will not disclose any of your individual information to managers of the HCA Directory or to any employer (including Alberta Health Services) unless at some later date you specifically request this in writing. Within the University, the research data may be audited by people external to the research team or by the Health Research Ethics Board, who would be bound by rules of confidentiality. Your results will remain confidential to the University team and your individual details will not appear in any report or publication.

 If you would like to participate please read the [information sheet](https://sites.ualberta.ca/~covidhca/info.html) then click Next, read and complete the consent form and if you are willing, move on to fill out the questionnaire.

 *Please note: If you are using a public computer, please aim to complete the survey in one sitting*

End of Block: Introduction

Start of Block: Consent Form 1

C1title **Consent Form**

C1header
 **The impact of Covid-19 on the health of Alberta Health Care Aides**
Principal Investigator: Dr. Nicola Cherry


 **CONSENT TO PARTICPATE IN THE STUDY**

C1p1 C1.1. Do you understand that you have been asked to be in a research study?

- Yes (1)
- No (2)

C1p2 C1.2. Have you read a copy of the [information sheet](https://sites.ualberta.ca/~covidhca/info.html)?

- Yes (1)
- No (2)

C1p3
C1.3. Do you understand the benefits and risks involved in taking part in this research study?
     *Note: There is a section on benefits and risks in the* [information sheet](https://sites.ualberta.ca/~covidhca/info.html)

- Yes (1)
- No (2)

C1p4
C1.4. Do you understand that you can phone 1–866–492–6093 or contact us at to ask questions or discuss the study?

- Yes (1)
- No (2)

C1p5
C1.5. Do you understand that you are free to withdraw from the study at any time, without having to give a reason and without affecting your employment or medical care?

- Yes (1)
- No (2)

C1p6
C1.6. Has the issue of confidentiality been explained to you?
    *Note: there is a paragraph on confidentiality in the* [information sheet](https://sites.ualberta.ca/~covidhca/info.html)*.*

- Yes (1)
- No (2)

C1p7
C1.7. Do you understand who will have access to your records?
    *Note: again, see the paragraph on confidentiality in the* [information sheet](https://sites.ualberta.ca/~covidhca/info.html)*.*

- Yes (1)
- No (2)

Display This Question:

If C1.1. Do you understand that you have been asked to be in a research study? = No

Or C1.2. Have you read a copy of the information sheet? = No

Or C1.3. Do you understand the benefits and risks involved in taking part in this research study?   ... = No

Or C1.4. Do you understand that you can phone 1–866–492–6093 or contact us at t... = No

Or C1.5. Do you understand that you are free to withdraw from the study at any time, without having... = No

Or C1.6. Has the issue of confidentiality been explained to you?     Note: there is a paragraph on c... = No

Or C1.7. Do you understand who will have access to your records?     Note: again, see the paragraph... = No

C1warning *You have checked 'no' to one of the above questions. If you would like to participate in this study please review your answers. Information about this study can be found in the* [information sheet](https://sites.ualberta.ca/~covidhca/info.html) *or by contacting us by phone (1-866-492-6093) or email.*

Display This Question:

If C1.1. Do you understand that you have been asked to be in a research study? = Yes

And C1.2. Have you read a copy of the information sheet? = Yes

And C1.3. Do you understand the benefits and risks involved in taking part in this research study?   ... = Yes

And C1.4. Do you understand that you can phone 1–866–492–6093 or contact us at t... = Yes

And C1.5. Do you understand that you are free to withdraw from the study at any time, without having... = Yes

And C1.6. Has the issue of confidentiality been explained to you?     Note: there is a paragraph on c... = Yes

And C1.7. Do you understand who will have access to your records?     Note: again, see the paragraph... = Yes

C1p8 C1.8. **If you agree** to take part in this research study, please type your name below and confirm today’s date.

Display This Question:

If C1.1. Do you understand that you have been asked to be in a research study? = Yes

And C1.2. Have you read a copy of the information sheet? = Yes

And C1.3. Do you understand the benefits and risks involved in taking part in this research study?   ... = Yes

And C1.4. Do you understand that you can phone 1–866–492–6093 or contact us at t... = Yes

And C1.5. Do you understand that you are free to withdraw from the study at any time, without having... = Yes

And C1.6. Has the issue of confidentiality been explained to you?     Note: there is a paragraph on c... = Yes

And C1.7. Do you understand who will have access to your records?     Note: again, see the paragraph... = Yes

C1p8_name
Name

________________________________________________________________

Display This Question:

If C1.1. Do you understand that you have been asked to be in a research study? = Yes

And C1.2. Have you read a copy of the information sheet? = Yes

And C1.3. Do you understand the benefits and risks involved in taking part in this research study?   ... = Yes

And C1.4. Do you understand that you can phone 1–866–492–6093 or contact us at t... = Yes

And C1.5. Do you understand that you are free to withdraw from the study at any time, without having... = Yes

And C1.6. Has the issue of confidentiality been explained to you?     Note: there is a paragraph on c... = Yes

And C1.7. Do you understand who will have access to your records?     Note: again, see the paragraph... = Yes

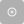

C1p8_date Date (day/month/year)

________________________________________________________________

End of Block: Consent Form 1

Start of Block: Consent Form 2

C2title **Consent to Linkage with health data in the Alberta administrative health database**

 By signing this consent you are agreeing to the study team collecting and using your personal health information from the Alberta administrative health database and allowing its inspection as described in the information sheet.

 **PLEASE NOTE: If you do not consent to this data linkage you are still most welcome to join the cohort and to complete the questionnaire today.**

C2p1

C2.1 I agree to Alberta Health linking my Alberta Health number to administrative health records and passing information to the investigators at the University of Alberta.

- Yes (1)
- No (2)

Display This Question:

If C2.1 I agree to Alberta Health linking my Alberta Health number to administrative health records... = Yes

C2p2
C2.2 Please type your name below and put today’s date

Display This Question:

If C2.1 I agree to Alberta Health linking my Alberta Health number to administrative health records... = Yes

C2p3name
Name

________________________________________________________________

Display This Question:

If C2.1 I agree to Alberta Health linking my Alberta Health number to administrative health records... = Yes

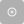

C2p3date
Date (day/month/year)

________________________________________________________________

Display This Question:

If C2.1 I agree to Alberta Health linking my Alberta Health number to administrative health records... = Yes

C2p4

 I know my Alberta Health Number

- Yes (1)
- No (2)

Display This Question:

If I know my Alberta Health Number   = Yes

C2p4number
 My Alberta Health number is:

________________________________________________________________

Display This Question:

If I know my Alberta Health Number   = Yes

C2p4dateofbirth
 My date of birth is (day/month/year):

________________________________________________________________

Display This Question:

If I know my Alberta Health Number   = No

C2p5

I agree to Alberta Health using my name, address and date of birth to find my Alberta Health Number.

- Yes (1)
- No (2)

Display This Question:

If I agree to Alberta Health using my name, address and date of birth to find my Alberta Health Numb... = Yes

C2p6
If you agree, please fill in the details below

- First name (1) __________________________________________________
- Middle name (if no middle name please type 'none') (2) __________________________________________________
- Last name (3) __________________________________________________

Display This Question:

If I agree to Alberta Health using my name, address and date of birth to find my Alberta Health Numb... = Yes

C2p7
 
 If you are not usually known by your first name, what is your preferred name?

________________________________________________________________

Display This Question:

If I agree to Alberta Health using my name, address and date of birth to find my Alberta Health Numb... = Yes

C2p8
 
What is your date of birth (day/month/year)?

________________________________________________________________

End of Block: Consent Form 2

Start of Block: Consent Form 3

C3title
**We plan to contact you again after the peak of the epidemic has passed.
 At that time we will ask for further consent, but if you are willing for us to contact you again please, type your name below and confirm today’s date.**

C3agree I agree to be contacted again.

- Yes (23)
- No (24)

Display This Question:

If I agree to be contacted again. = Yes

C3name Name

________________________________________________________________

Display This Question:

If I agree to be contacted again. = Yes

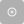

C3date Date (day/month/year)

________________________________________________________________

Display This Question:

If I agree to be contacted again. = Yes

C3subtitle Please give your email and phone number below, so that we can contact you again.

Display This Question:

If I agree to be contacted again. = Yes

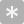

C3email Email

________________________________________________________________

Display This Question:

If I agree to be contacted again. = Yes

C3phone Phone number

________________________________________________________________

End of Block: Consent Form 3

Start of Block: Demographics

Qa
**Thank you for agreeing to fill in the questionnaire. It should not take too long.**

**May we first collect a few demographic details**

**A) Demographics**

A.1 Which gender do you identify with?

- Male (11)
- Female (12)
- Other (13)

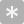

A.2 What is your age in years (today)?

________________________________________________________________

A.3 Are you now

- Married, or living as married? (1)
- Widowed, divorced? (2)
- Single? (3)

A.4 Do you have any children <18 years living in your household?

- Yes (1)
- No (2)

A.5 Do you have a Health Care Aide certificate?

- Yes (23)
- No (24)

Display This Question:

If Do you have a Health Care Aide certificate? = Yes

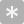

A.6 In what year did you get your HCA certificate?

________________________________________________________________

End of Block: Demographics

Start of Block: Introduction

Introduction **The rest of this questionnaire is about the period since March 6th – the day the first Covid-19 case was diagnosed in Alberta.**
 **When we ask about your work in a HCA role we mean work for which, in Alberta, you needed to be on the HCA register.**

End of Block: Introduction

Start of Block: Block 1

Q101 **B) Employment since March 6th**

Q1.0 Since March 6th 2020, have you been carrying out ***any*** role as a HCA in Alberta?

- Yes (23)
- No (24)

Display This Question:

If Since March 6th 2020, have you been carrying out any role as a HCA in Alberta? = No

| 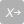 | 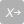 |
| --- | --- |

Q1.1 Why not?

- Retired (1)
- Sickness/disability with onset before 6th March (2)
- Maternity leave (3)
- Family responsibilities (4)
- Other, namely (5) __________________________________________________

End of Block: Block 1

Start of Block: Block 2

Q2.0 Since 6th March 2020 have you had any period when you have been self-isolating (including quarantine) at home and unable to attend work ***because of you were at high risk to spread the Covid-19 virus?***

- Yes (23)
- No (24)

Display This Question:

If Since 6th March 2020 have you had any period when you have been self-isolating (including quarant... = Yes

| 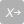 | 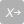 |
| --- | --- |

Q2.1 Why was this?

- You had an infection or symptoms known, presumed or suspected to be caused by Covid-19 (1)
- A family member had an infection or symptoms known, presumed or suspected to be caused by Covid-19 (2)
- You had (inadequately protected) contact with a patient or colleague with known, presumed or suspected Covid-19 (3)
- You had returned from international travel (4)
- Other, namely (5) __________________________________________________

Display This Question:

If Since 6th March 2020 have you had any period when you have been self-isolating (including quarant... = Yes

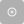

Q2.2 What was the date of the first day of this self-isolation?

- Day (8) __________________________________________________
- Month (9) __________________________________________________
- Year (10) __________________________________________________

Display This Question:

If Since 6th March 2020 have you had any period when you have been self-isolating (including quarant... = Yes

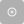

Q2.3 What was (or will be) the date of the last day of this self-isolation?

- Day (8) __________________________________________________
- Month (9) __________________________________________________
- Year (10) __________________________________________________

End of Block: Block 2

Start of Block: Block 3

Q3.0 Since 6th March 2020 have you had any period when you have been self-isolating at home but **NOT** because you were at high risk to spread COVID-19?

- Yes (23)
- No (24)

Display This Question:

If Since 6th March 2020 have you had any period when you have been self-isolating at home but NOT be... = Yes

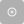

Q3.1 What was the date of the first day of this self-isolation?

- Day (8) __________________________________________________
- Month (9) __________________________________________________
- Year (10) __________________________________________________

Display This Question:

If Since 6th March 2020 have you had any period when you have been self-isolating at home but NOT be... = Yes

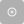

Q3.2 What was (or will be) the date of the last day of this self-isolation?

- Day (8) __________________________________________________
- Month (9) __________________________________________________
- Year (10) __________________________________________________

End of Block: Block 3

Start of Block: Block 4

Q4.0 In the most recent week you have worked since March 6th, has your role as a HCA in Alberta involved **contact with patients or clients?**

- Yes (23)
- No (24)

Display This Question:

If In the most recent week you have worked since March 6th, has your role as a HCA in Alberta involv... = Yes

| 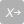 | 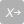 |
| --- | --- |

Q4.1 What role have you held while in contact with patients or clients? (check all that apply)

- HCA working in an inpatient setting in a hospital (1)
- HCA in ambulatory or outpatient settings in a hospital (2)
- HCA in an emergency room (3)
- HCA in a community health setting (4)
- HCA providing clinical support to a residential institution or contained community (such as care home, prison, first nations), please specify (5) __________________________________________________
- HCA providing clinical support to a workforce, please specify (6) __________________________________________________
- Other HCA role, please specify (7) __________________________________________________

Display This Question:

If What role have you held while in contact with patients or clients? (check all that apply) = HCA working in an inpatient setting in a hospital

| 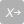 | 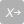 |
| --- | --- |

Q4.1.1.1 In the last week you have worked since 6th March were you working in (Check all that apply)

- Critical care (1)
- ICU (2)
- Wards designated for the care of infectious patients (3)
- General (adult) medical wards (4)
- Wards for the care of geriatric patients (5)
- Wards for the care of psychiatric patients (6)
- Wards for the care of pediatric patients	(7)
- Wards for the care of obstetrical patients (8)
- Other, namely (8) __________________________________________________

Display This Question:

If In the most recent week you have worked since March 6th, has your role as a HCA in Alberta involv... = No

| 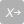 | 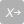 |
| --- | --- |

Q4.2 In the most recent week you have worked since March 6th 2020, was your role as a HCA primarily without contact with patients or clients because you were working in

- Public health? (1)
- Administration? (2)
- Teaching or research? (3)
- Other, namely (4) __________________________________________________

End of Block: Block 4

Start of Block: Block 5

Display This Question:

If In the most recent week you have worked since March 6th, has your role as a HCA in Alberta involv... = Yes

| 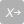 | 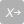 |
| --- | --- |

Q5.0 What was the age range of your patients in your most recent week at work since March 6th? Please check one or more below to reflect the great majority of your patients

- All ages including children (1)
- All ages 18 years and older (2)
- (3)
- 5 (4)
- 18 (5)
- 55 (6)
- >=70 years? (7)
- Other, namely (8) __________________________________________________

End of Block: Block 5

Start of Block: Block 6

Q6.0 ***In your most recent week at work since March 6th,*** how many days did you work in a HCA role in Alberta?

________________________________________________________________

Q6title ***In your most recent week at work since March 6th,***

Q6.1 What was the **total number of hours in that week** you worked and had **in person** contact with patients or clients? (best estimate)

________________________________________________________________

Q6.1.1 **How many patients or clients did you see in person** in your most recent week at work since March 6th? (best estimate)

________________________________________________________________

Q6.1.2 What proportion of the patients or clients you saw **in person** in that week were screened (for symptoms or fever) before you were in contact with them?

 **if no patients seen in person please check 100%**

- 100% (1)
- >90% but not 100% (2)
- 50-90% (3)
- some, but	(4)
- none (5)

Q6.2 What was the total number of hours in that week you worked with patients or clients by **phone, email or videoconferencing**? (best estimate)

________________________________________________________________

Q6.3 What was the total number of hours in that week you worked as a HCA, without patient or client contact? (best estimate)

________________________________________________________________

End of Block: Block 6

Start of Block: Block 7

Q7.0 Since March 6th has your work involved contact with known, presumed or suspected Covid-19 patients or clients?

- Yes (25)
- No (26)
- Don't know (29)

Skip To: End of Block If Since March 6th has your work involved contact with known, presumed or suspected Covid-19 patient... != Yes

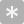

Q7.1
Since March 6th on how many days have you had one-on-one contact with known, presumed or suspected COVID-19 patients or clients?

________________________________________________________________

Q7.2 Since March 6th, have you taken part in (or been within 2 m of) aerosol generating medical procedures (see below)?

- Yes (23)
- No (24)

Display This Question:

If Since March 6th, have you taken part in (or been within 2 m of) aerosol generating medical proced... != No

| 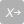 | 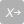 |
| --- | --- |

Q7.2.a If yes, what procedures? (check all that apply)

- Intubation and related procedures (manual ventilation, open endotracheal suctioning, extubation) (1)
- Cardiopulmonary resuscitation (2)
- Bi-level Positive Airway Pressure (BiPAP, CPAP) (3)
- Humidified high flow oxygen systems (ARVO, Optiflow) (4)
- Tracheostomy care (5)
- Bronchoscopy (6)
- Sputum induction (7)
- Nebulized/aerosolized medication administration (8)
- Open respiratory/airway suctioning (9)
- High frequency oscillatory ventilation (10)
- Other, namely (11) __________________________________________________

| Page Break |
| --- |

Display This Question:

If Since March 6th, have you taken part in (or been within 2 m of) aerosol generating medical proced... = Yes

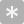

Q7.3 Please estimate on how many days since 6th March you have been involved in aerosol generating medical procedures

________________________________________________________________

Display This Question:

If Since March 6th, have you taken part in (or been within 2 m of) aerosol generating medical proced... = Yes

Q7.4 Were any of these on patients or clients with known, presumed or suspected Covid-19?

- Yes (25)
- No (26)
- Don't know (27)

| Page Break |
| --- |

Q7.5 Have there been any times when you have had contact with known, presumed or suspected COVID-19 patients or clients when you, personally, did NOT HAVE ACCESS to all the approved Personal Protective Equipment (PPE)?

- Yes (23)
- No (24)

Display This Question:

If Have there been any times when you have had contact with known, presumed or suspected COVID-19 pa... = Yes

Q7.5.1 Which types of (clean) PPE were not available?

- N 95 respirator (4)
- Procedure mask (7)
- Eye protection (8)
- Face shield (9)
- Gown (10)
- Gloves (11)
- Other, namely (12) __________________________________________________

Q7.6 Have there been any times when you have had contact with known, presumed or suspected COVID-19 patients or clients where appropriate PPE was available but you did not use it effectively?

- Yes (23)
- No (24)

Display This Question:

If Have there been any times when you have had contact with known, presumed or suspected COVID-19 pa... = Yes

Q7.6.1 Please describe the incident where PPE was not used effectively.

________________________________________________________________

________________________________________________________________

________________________________________________________________

________________________________________________________________

________________________________________________________________

End of Block: Block 7

Start of Block: Block 8

Qc **C) Your perceptions and concerns**

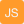

Q8.1 Please mark on the line below **how true** the following statements are about your work, during your most recent week at work since March 6th, compared with working before March 6th.

|  | **Completely disagree** | **Completely agree** |
| --- | --- | --- |

| My hours of work are about the same (1) |
| --- |
| My work tasks are about the same (4) |
| The patient or client make-up is about the same (5) |
| My patients or clients are no more stressed (6) |
| My co-workers are no more stressed (7) |
| I am no more stressed (8) |

| Page Break |
| --- |

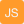

Q8.2 Please mark on the line below **how confident** you feel about working with patients or clients with known, presumed or suspected COVID-19.

|  | **Not at all confident** | **Very confident** |
| --- | --- | --- |

| I have the clinical knowledge I need (1) |
| --- |
| I know how to use the required PPE (9) |
| I know how and when to refer for testing (10) |
| I know how and when to refer for treatment (14) |
| I have the support I need from co-workers (15) |
| I have access to all the required PPE (16) |
| There are sufficient staff to do the job safely (17) |

| Page Break |
| --- |

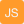

Q8.3 Please mark on the line below to **show your worries** about the COVID-19 epidemic.

|  | **Not at all worried** | **Very worried** |
| --- | --- | --- |

| That I shall be infected (1) |
| --- |
| That I shall infect my family (21) |
| That I shall infect my patients (22) |
| That I shall infect my co-workers/colleagues (23) |
| That I shall not be able to cope with the work (24) |
| That I shall have to let people die (25) |
| That my experience is inadequate (26) |
| That I shall fail myself and my family (27) |

| Page Break |
| --- |

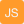

Q8.4 Please mark on the line below to show where you will **find support** during this time.

|  | **No support at all** | **Very strong support** |
| --- | --- | --- |

| My immediate family (1) |
| --- |
| My colleagues or co-workers (19) |
| A senior colleague or mentor (20) |
| My immediate organization (21) |
| Alberta Health Services (23) |
| Alberta's Chief Medical Officer of Health (24) |
| The managers of the HCA registry (27) |
| My religious community (28) |

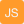

Q8.4.5

|  | **No support at all** | **Very strong support** |
| --- | --- | --- |

| Others, namely (1) |
| --- |

End of Block: Block 8

Start of Block: Block 9

Q9title Considering events affecting your work as a HCA during the COVID-19 epidemic (since 6th March),

Q9.1 What has been the most difficult or stressful event you have had to deal with?
 *(If you prefer not to answer, please put "N/A" and go to the next question)*

________________________________________________________________

________________________________________________________________

________________________________________________________________

________________________________________________________________

________________________________________________________________

Q9.2 What has been the event that has most reinforced your pride in your professional behaviour?
 *(If you prefer not to answer, please put "N/A" and go to the next question)*

________________________________________________________________

________________________________________________________________

________________________________________________________________

________________________________________________________________

________________________________________________________________

End of Block: Block 9

Start of Block: Block 10

Q10.0 Do you have reason to believe that you may have been infected with the COVID-19 virus?

- Yes (23)
- No (24)

Skip To: End of Block If Do you have reason to believe that you may have been infected with the COVID-19 virus? = No

Q10.1 Have you been tested for COVID-19?

- Yes (23)
- No (24)

Skip To: End of Block If Have you been tested for COVID-19? = No

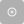

Q10.2 What date were you tested?

- Day (1) __________________________________________________
- Month (2) __________________________________________________
- Year (3) __________________________________________________

Q10.3 Have you had the result?

- Yes (23)
- No (24)

Display This Question:

If Have you had the result? = Yes

Q10.4 What was the result?

- Positive (4)
- Negative (5)

End of Block: Block 10

Start of Block: Bridge

Q11title
**Your Health**
  Next, we would ask you to complete two short questionnaires, one on symptoms you may have experienced for at least TWO CONSECUTIVE DAYS since March 6th. and then a short mood scale to tell us how you have been coping in the last week.

End of Block: Bridge

Start of Block: Block 11

Q11.0 Since March 6th, have you had an episode when you have been unwell for two or more consecutive days (whether or not you reported for duty)?

- Yes (23)
- No (24)

Skip To: End of Block If Since March 6th, have you had an episode when you have been unwell for two or more consecutive da... = No

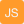

Q11.1 Please mark below **how much you have been bothered** FOR TWO OR MORE CONSECUTIVE DAYS since March 6th for each symptom/problem listed below. If more than one incident, record the symptoms/problems from the one that bothered you most.

|  | Not at all bothered | A little | Moderately | Quite a bit | Extremely bothered |
| --- | --- | --- | --- | --- | --- |

| Coughing (1) |
| --- |
| Chest pains (4) |
| Coughing up sputum (5) |
| Coughing up blood (6) |
| Sweating (7) |
| Chills (8) |
| Headache (9) |
| Nausea (10) |
| Vomiting (11) |
| Diarrhea (12) |
| Stomach pain (13) |
| Muscle pain (14) |
| Lack of appetite (15) |
| Trouble concentrating (16) |
| Trouble thinking (17) |
| Trouble sleeping (18) |
| Fatigue (19) |

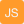

Q11.17

|  | Not at all bothered | A little | Moderately | Quite a bit | Extremely bothered |
| --- | --- | --- | --- | --- | --- |

| Other, namely (1) |
| --- |

End of Block: Block 11

Start of Block: Block 12

Q12title
In the mood questionnaire below, please check the box alongside the reply that is closest to how you have been feeling IN THE PAST WEEK. **Don't take too long over your replies; your immediate answer is best.**

Q12.1
*In the past week* I feel tense or 'wound up':

- Most of the time (1)
- A lot of the time (2)
- From time to time, occasionally (3)
- Not at all (4)

Q12.2 *In the past week*
 I still enjoy the things I used to enjoy:

- Definitely as much (1)
- Not quite so much (2)
- Only a little (3)
- Hardly at all (4)

Q12.3 *In the past week*
 I get a sort of frightened feeling as if something awful is about to happen:

- Very definitely and quite badly (1)
- Yes, but not too badly (2)
- A little but it doesn't worry me (3)
- Not at all (4)

Q12.4 *In the past week*
 I can laugh and see the funny side of things:

- As much as I always could (1)
- Not quite so much now (2)
- Definitely not so much now (3)
- Not at all (4)

Q12.5 *In the past week*
 Worrying thoughts go through my mind:

- A great deal of the time (1)
- A lot of the time (2)
- Not too often (3)
- Very little (4)

Q12.6 *In the past week*
 I feel cheerful:

- Never (1)
- Not often (2)
- Sometimes (3)
- Most of the time (4)

Q12.7 *In the past week*
 I can sit at ease and feel relaxed:

- Definitely (1)
- Usually (2)
- Not Often (3)
- Not at all (4)

Q12.8 *In the past week*
 I feel as if I am slowed down:

- Nearly all the time (1)
- Very often (2)
- Sometimes (3)
- Not at all (4)

Q12.9 *In the past week*
 I get a sort of frightened feeling like ‘butterflies’ in the stomach:

- Not at all (1)
- Occasionally (2)
- Quite often (3)
- Very often (4)

Q12.10 *In the past week*
 I have lost interest in my appearance:

- Definitely (1)
- I don’t take as much care as I should (2)
- I may not take quite as much care (3)
- I take just as much care as ever (4)

Q12.11 *In the past week*
 I feel restless as if I have to be on the move:

- Very much indeed (1)
- Quite a lot (2)
- Not very much (3)
- Not at all (4)

Q12.12 *In the past week*
 I look forward with enjoyment to things:

- As much as I ever did (1)
- Rather less than I used to (2)
- Definitely less than I used to (3)
- Hardly at all (4)

Q12.13 *In the past week*
 I get sudden feelings of panic:

- Very often indeed (1)
- Quite often (2)
- Not very often (3)
- Not at all (4)

Q12.14 *In the past week*
 I can enjoy a good book or radio or television program:

- Often (1)
- Sometimes (2)
- Not often (3)
- Very seldom (4)

End of Block: Block 12

Start of Block: Block 13

Q13.1 **Has a physician ever told you that you had asthma?**

- Yes (1)
- No (2)

Display This Question:

If Has a physician ever told you that you had asthma? = Yes

Q13.1.1 In the 12 months up to March 6th did you have any asthma symptoms or asthma attacks?

- Yes (1)
- No (2)

Display This Question:

If Has a physician ever told you that you had asthma? = Yes

Q13.1.2 In the 12 months up to March 6th, have you taken any medicine for asthma such as inhalers (pumps), nebulizers, pills, liquids or injections?

- Yes (1)
- No (2)

Q13.2 **Do you have chronic bronchitis, emphysema or chronic obstructive pulmonary disease or COPD?**

- Yes (1)
- No (2)

Q13.3 **Have you ever been treated for anxiety or depression?** In ‘treatment’ we include both medication and psychological interventions.

- Yes (1)
- No (2)

Display This Question:

If Have you ever been treated for anxiety or depression? In ‘treatment’ we include both medication a... = Yes

Q13.3.1 In the 12 months up to March 6th, did you receive any treatment for anxiety?

- Yes (1)
- No (2)

Display This Question:

If Have you ever been treated for anxiety or depression? In ‘treatment’ we include both medication a... = Yes

Q13.3.2 In the 12 months up to March 6th, did you receive any treatment for depression?

- Yes (1)
- No (2)

| Page Break |
| --- |

Q13.4 In the 12 months up to March 6th, did you have any other chronic condition?

- Yes (1)
- No (2)

Display This Question:

If In the 12 months up to March 6th, did you have any other chronic condition? = Yes

Q13.4.1 If yes, please specify

________________________________________________________________

| Page Break |
| --- |

Q13.5 Have you ever smoked at least one cigarette a day for as long as a year?

- Yes (1)
- No (2)

Display This Question:

If Have you ever smoked at least one cigarette a day for as long as a year? = Yes

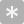

Q13.5.1 If yes, at what age did you start?

________________________________________________________________

Display This Question:

If Have you ever smoked at least one cigarette a day for as long as a year? = Yes

Q13.5.2 Did you smoke cigarettes in the 12 months leading up to March 6th?

- Yes (4)
- No (5)

Display This Question:

If Have you ever smoked at least one cigarette a day for as long as a year? = Yes

And Did you smoke cigarettes in the 12 months leading up to March 6th? = No

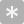

Q13.5.3 If no, what age did you stop?

________________________________________________________________

Display This Question:

If Have you ever smoked at least one cigarette a day for as long as a year? = Yes

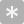

Q13.5.4 How many cigarettes a day do you/did you smoke?

________________________________________________________________

End of Block: Block 13

Start of Block: Block 14

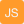

**Cohort profile: Recruitment and retention in a prospective cohort of Canadian health care workers during the Covid-19 pandemic.**

Nicola Cherry, Anil Adisesh, Igor Burstyn, Quentin Durand-Moreau, Jean-Michel Galarneau, France Labreche, Shannon Ruzycki, Tanis Zadunayski

Supplementary materials 2: additional tables

Table SM 1: Participants giving a pre-vaccine serology sample by baseline characteristics

Table SM 2: Participants giving at least one post-vaccine serology sample by baseline characteristics: participants reporting one or more vaccine doses

Table SM 1: Participants giving a pre-vaccine serology sample by baseline characteristics

|  |  | Pre-vaccine sample given | | | Bivariate | | | Multivariable* | | |
| --- | --- | --- | --- | --- | --- | --- | --- | --- | --- | --- |
| Factor |  | n | % | N | OR | 95% CI | p= | OR | 95% CI | p= |
| Baseline questionnaire completed | Short or partial | 231 | 38.6 | 598 | ‒ | ‒ | ‒ | ‒ | ‒ | ‒ |
|  | Fully | 2680 | 62.4 | 4297 | 2.63 | 2.21-3.14 | <0.001 | ‒ | ‒ | ‒ |
|  | Fully but not working | 29 | 42.0 | 69 | 1.15 | 0.69-1.91 | 0.584 | ‒ | ‒ | ‒ |
| Gender | Not female | 470 | 55.1 | 853 | 1 | ‒ | ‒ | 1 | ‒ | ‒ |
|  | Female | 2470 | 60.1 | 4111 | 1.23 | 1.06-1.42 | 0.007 | 1.27 | 1.05-1.53 | 0.012 |
| Age | < 40 y | 1040 | 55.8 | 1863 | 1 | ‒ | ‒ | 1 | ‒ | ‒ |
|  | 40 < 55 y | 1091 | 59.3 | 1839 | 1.15 | 1.01-1.32 | 0.031 | 1.30 | 1.12-1.51 | 0.001 |
|  | 55+ | 809 | 64.1 | 1262 | 1.41 | 1.22-1.64 | <0.001 | 1.63 | 1.36-1.95 | <0.001 |
| Work role** | MD | 836 | 58.0 | 1442 | 1 | ‒ | ‒ | 1 | ‒ | ‒ |
|  | RN | 1993 | 63.6 | 3136 | 1.26 | 1.11-1.44 | <0.001 | 1.38 | 1.18-1.61 | <0.001 |
|  | LPN | 38 | 53.5 | 71 | 0.83 | 0.52-1.35 | 0.459 | 0.78 | 0.46-1.33 | 0.355 |
|  | PSW | 38 | 16.2 | 235 | 0.14 | 0.10-0.20 | <0.001 | 0.14 | 0.09-0.22 | <0.001 |
|  | HCA | 35 | 43.8 | 80 | 0.56 | 0.36-0.89 | 0.013 | 0.68 | 0.40-1.13 | 0.133 |
| Married or de-facto | No | 678 | 56.5 | 1200 | 1 | ‒ | ‒ | 1 | ‒ | ‒ |
|  | Yes | 2204 | 60.8 | 3623 | 1.20 | 1.05-1.36 | 0.008 | 1.26 | 1.07-1.47 | 0.005 |
|  | Unknown | 58 | 41.1 | 141 | 0.54 | 0.38-0.77 | 0.001 | ‒ | ‒ | ‒ |
| Child at home < 18 y | No | 1707 | 60.2 | 2835 | 1 | ‒ | ‒ | 1 | ‒ | ‒ |
|  | Yes | 1225 | 58.1 | 2107 | 0.92 | 0.82-1.03 | 0.143 | 0.86 | 0.74-1.00 | 0.047 |
|  | Unknown | 8 | 36.4 | 22 | 0.38 | 0.16-0.90 | 0.029 | ‒ | ‒ | ‒ |
| Smoked tobacco in last 12 months | No | 2615 | 63.4 | 4127 | 1 | ‒ | ‒ | 1 | ‒ | ‒ |
|  | Yes | 126 | 46.3 | 272 | 0.50 | 0.39-0.64 | <0.001 | 0.62 | 0.47-0.82 | 0.001 |
|  | Unknown | 199 | 35.2 | 565 | 0.31 | 0.26-0.38 | <0.001 | ‒ | ‒ | ‒ |
| Chronic lung disease | No | 2694 | 62.3 | 4127 | 1 | ‒ | ‒ | 1 | ‒ | ‒ |
|  | Yes | 47 | 60.3 | 272 | 0.92 | 0.58-1.45 | 0.714 | 1.21 | 0.70-2.09 | 0.492 |
|  | Unknown | 199 | 35.5 | 565 | 0.33 | 0.28-0.40 | <0.001 | ‒ | ‒ | ‒ |
| Asthma medications in last 12 months | No | 2326 | 61.4 | 3787 | 1 | ‒ | ‒ | 1 | ‒ | ‒ |
|  | Yes | 415 | 67.3 | 617 | 1.29 | 1.08-1.55 | 0.006 | 1.31 | 1.08-1.58 | 0.006 |
|  | Unknown | 199 | 35.5 | 560 | 0.35 | 0.29-0.42 | <0.001 | ‒ | ‒ | ‒ |
| Treatment for anxiety/depression in last 12 months | No | 2082 | 62.0 | 3357 | 1 | ‒ | ‒ | 1 | ‒ | ‒ |
|  | Yes | 659 | 63.0 | 1046 | 1.04 | 0.90-1.20 | 0.567 | 1.08 | 0.93-1.26 | 0.328 |
|  | Unknown | 199 | 35.5 | 561 | 0.34 | 0.28-0.41 | <0.001 | ‒ | ‒ | ‒ |
| Working directly with patients | No | 348 | 60.4 | 576 | 1 | ‒ | ‒ | 1 | ‒ | ‒ |
|  | Yes | 2513 | 60.1 | 4184 | 0.99 | 0.82-1.18 | 0.871 | 1.19 | 0.97-1.45 | 0.092 |
|  | Unknown | 79 | 38.7 | 204 | 0.41 | 0.30-0.57 | <0.001 | ‒ | ‒ | ‒ |
|  | N | 2940 | 59.2 | 4964 | 4964 |  |  | 4282* |  |  |

*Restricted to those who completed the full baseline questionnaire

**MD, medical doctor; RN registered nurse or registered psychiatric nurse; LPN, licensed practical nurse; PSW, personal support worker; HCA, health care aide.

Table SM2: Participants giving at least one post-vaccine serology sample by baseline characteristics: participants reporting one or more vaccine doses

|  |  | Post-vaccine sample given | | | Bivariate | | | Multivariable* | | |
| --- | --- | --- | --- | --- | --- | --- | --- | --- | --- | --- |
| Factor |  | n | % | N | OR | 95% CI | p= | OR | 95% CI | p= |
| Baseline questionnaire completed | Short or partial | 223 | 44.7 | 499 | 1 | ‒ | ‒ | ‒ | ‒ | ‒ |
|  | Fully | 2502 | 62.3 | 4015 | 2.05 | 1.70-2.47 | <0.001 | ‒ | ‒ | ‒ |
|  | Fully but not working | 27 | 50.9 | 53 | 1.29 | 0.73-2.27 | 0.385 | ‒ | ‒ | ‒ |
| Gender | Not female | 460 | 58.0 | 793 | 1 | ‒ | ‒ | 1 | ‒ | ‒ |
|  | Female | 2292 | 60.7 | 3774 | 1.12 | 0.96-1.31 | 0.154 | 1.20 | 0.99-1.45 | 0.068 |
| Age | < 40 y | 925 | 54.1 | 1709 | 1 | ‒ | ‒ | 1 | ‒ | ‒ |
|  | 40 < 55 y | 1038 | 60.7 | 1711 | 1.31 | 1.14-1.50 | <0.001 | 1.50 | 1.28-1.75 | <0.001 |
|  | 55+ | 789 | 68.8 | 1147 | 1.87 | 1.60-2.19 | <0.001 | 2.10 | 1.74-2.54 | <0.001 |
| Work role** | MD | 839 | 61.1 | 1374 | 1 | ‒ | ‒ | 1 | ‒ | ‒ |
|  | RN | 1830 | 63.3 | 2889 | 1.10 | 0.97-1.26 | 0.150 | 1.18 | 1.01-1.39 | 0.041 |
|  | LPN | 34 | 50.7 | 67 | 0.66 | 0.40-1.07 | 0.094 | 0.62 | 0.36-1.07 | 0.086 |
|  | PSW | 20 | 11.6 | 173 | 0.08 | 0.05-0.13 | <0.001 | 0.06 | 0.03-0.10 | <0.001 |
|  | HCA | 29 | 45.3 | 64 | 0.53 | 0.32-0.87 | 0.013 | 0.59 | 0.34-1.02 | 0.061 |
| Married or de-facto | No | 613 | 57.0 | 1076 | 1 | ‒ | ‒ | 1 | ‒ | ‒ |
|  | Yes | 2088 | 62.0 | 3369 | 1.23 | 1.07-1.42 | 0.003 | 1.25 | 1.06-1.48 | 0.008 |
|  | Unknown | 51 | 41.8 | 122 | 0.54 | 0.37-0.79 | 0.002 | ‒ | ‒ | ‒ |
| Child at home < 18 y | No | 1601 | 62.1 | 2579 | 1 | ‒ | ‒ | 1 | ‒ | ‒ |
|  | Yes | 1144 | 58.1 | 1969 | 0.85 | 0.75-0.95 | 0.007 | 0.81 | 0.70-0.95 | 0.009 |
|  | Unknown | 7 | 36.8 | 19 | 0.36 | 0.14-0.91 | 0.031 | ‒ | ‒ | ‒ |
| Smoked tobacco in last 12 months | No | 2446 | 63.1 | 3876 | 1 | ‒ | ‒ | 1 | ‒ | ‒ |
|  | Yes | 112 | 49.3 | 227 | 0.57 | 0.44-0.74 | <0.001 | 0.76 | 0.56-1.04 | 0.083 |
|  | Unknown | 194 | 41.8 | 464 | 0.42 | 0.35-0.51 | <0.001 | ‒ | ‒ | ‒ |
| Chronic lung disease | No | 2523 | 62.5 | 4037 | 1 | ‒ | ‒ | 1 | ‒ | ‒ |
|  | Yes | 37 | 52.9 | 70 | 0.67 | 0.42-1.08 | 0.101 | 0.63 | 0.37-1.09 | 0.102 |
|  | Unknown | 192 | 41.7 | 460 | 0.43 | 0.35-0.52 | <0.001 | ‒ | ‒ | ‒ |
| Asthma medications in last 12 months | No | 2181 | 61.9 | 3526 | 1 | ‒ | ‒ | 1 | ‒ | ‒ |
|  | Yes | 379 | 65.1 | 582 | 1.15 | 0.96-1.38 | 0.132 | 1.17 | 0.96-1.42 | 0.114 |
|  | Unknown | 192 | 41.8 | 459 | 0.44 | 0.36-0.54 | <0.001 | ‒ | ‒ | ‒ |
| Treatment for anxiety/depression in last 12 months | No | 1958 | 62.5 | 3135 | 1 | ‒ | ‒ | 1 | ‒ | ‒ |
|  | Yes | 602 | 61.9 | 972 | 0.98 | 0.84-1.13 | 0.769 | 1.05 | 0.89-1.23 | 0.555 |
|  | Unknown | 192 | 41.7 | 460 | 0.43 | 0.35-0.53 | <0.001 | ‒ | ‒ | ‒ |
| Working directly with patients | No | 313 | 60.8 | 515 | 1 | ‒ | ‒ | 1 | ‒ | ‒ |
|  | Yes | 2362 | 60.8 | 3884 | 1.00 | 0.83-1.21 | 0.987 | 1.28 | 1.04-1.57 | 0.021 |
|  | Unknown | 77 | 45.8 | 168 | 0.55 | 0.38-0.78 | 0.001 | ‒ | ‒ | ‒ |
| Total | N | 2752 |  | 4567 | 4567 | ‒ | ‒ | 4004* | ‒ | ‒ |

* Only these who completed the full baseline questionnaire and had received at least one vaccine dose

**MD, medical doctor; RN registered nurse or registered psychiatric nurse; LPN, licensed practical nurse; PSW, personal support worker; HCA, health care aide.
